# LD-informed deep learning for Alzheimer’s gene loci detection using WGS data

**DOI:** 10.1101/2024.09.19.24313993

**Authors:** Taeho Jo, Paula Bice, Kwangsik Nho, Andrew J. Saykin, the Alzheimer’s Disease Sequencing Project

**Author notes:** **Corresponding authors** Andrew J. Saykin, PsyD* Indiana Alzheimer Disease Research Center and Center for Neuroimaging, Department of Radiology and Imaging Sciences, Indiana University School of Medicine, Indianapolis, IN 46202, USA ’ Kwangsik Nho, PhD* Indiana Alzheimer Disease Research Center and Center for Neuroimaging, Department of Radiology and Imaging Sciences, Indiana University School of Medicine, Indianapolis, IN 46202, USA Taeho Jo, PhD* Indiana Alzheimer Disease Research Center and Center for Neuroimaging, Department of Radiology and Imaging Sciences, Indiana University School of Medicine, Indianapolis, IN 46202, USA.

## Abstract

**INTRODUCTION:** The exponential growth of genomic datasets necessitates advanced analytical tools to effectively identify genetic loci from large-scale high throughput sequencing data. This study presents Deep-Block, a multi-stage deep learning framework that incorporates biological knowledge into its AI architecture to identify genetic regions as significantly associated with Alzheimer’s disease (AD). The framework employs a three-stage approach: (1) genome segmentation based on linkage disequilibrium (LD) patterns, (2) selection of relevant LD blocks using sparse attention mechanisms, and (3) application of TabNet and Random Forest algorithms to quantify single nucleotide polymorphism (SNP) feature importance, thereby identifying genetic factors contributing to AD risk.

**METHODS:** The Deep-Block was applied to a large-scale whole genome sequencing (WGS) dataset from the Alzheimer’s Disease Sequencing Project (ADSP), comprising 7,416 non-Hispanic white participants (3,150 cognitively normal older adults (CN), 4,266 AD).

**RESULTS:** 30,218 LD blocks were identified and then ranked based on their relevance with Alzheimer’s disease. Subsequently, the Deep-Block identified novel SNPs within the top 1,500 LD blocks and confirmed previously known variants, including *APOE* rs429358 and rs769449. Expression Quantitative Trait Loci (eQTL) analysis across 13 brain regions provided functional evidence for the identified variants. The results were cross-validated against established AD-associated loci from the European Alzheimer’s and Dementia Biobank (EADB) and the GWAS catalog.

**DISCUSSION:** The Deep-Block framework effectively processes large-scale high throughput sequencing data while preserving SNP interactions during dimensionality reduction, minimizing bias and information loss. The framework’s findings are supported by tissue-specific eQTL evidence across brain regions, indicating the functional relevance of the identified variants. Additionally, the Deep-Block approach has identified both known and novel genetic variants, enhancing our understanding of the genetic architecture and demonstrating its potential for application in large-scale sequencing studies.

## 1 Introduction

The advancement of deep learning in artificial intelligence has introduced new frameworks for analyzing complex genetic inheritance patterns, enhancing the interpretation of genomic data ^1–4^. For complex diseases such as Alzheimer’s disease (AD), there is a critical need for advanced analytic tools provided by Artificial Intelligence (AI) to decipher the complexities of human genetic makeup ^4–6^. The complexity of genomic studies necessitates innovative and adaptive approaches that transcend traditional machine learning techniques to analyze and elucidate these intricate genetic interactions ^7, 8^. The high dimensionality and large sample sizes characteristic of genetic data in AD research underscore the necessity for methods capable of navigating the complex landscape ^9–11^. While several machine learning-based dimensionality reduction methods have been proposed, they have encountered challenges such as loss of phenotypic association information during the reduction process, reproducibility issues, and data-dependent inconsistencies in results ^12–14^.

Here, we present Deep-Block, a deep learning framework designed to address the complexities of genomic sequencing data through targeted analysis of whole genome sequencing (WGS) data. Deep-Block employs a linkage disequilibrium (LD) block-based approach to systematically identify significant genetic regions, aiming to preserve vital phenotypic associations and minimize the loss of genetic information crucial for understanding disease phenotypes. The framework incorporates advanced machine learning and genomic imputation techniques ^15–17^ to ensure a comprehensive dataset without any missing values for analysis. Furthermore, the integration of the TabNet model ^18, 19^, an attention-based neural network, enhances the process by providing a detailed assessment of feature importance within the genetic data, thus enriching the analysis. The calculation of Phenotype Influence Scores (PIS) offers additional insights into the genetic basis of the disease, informing future research directions. The framework includes Expression Quantitative Trait Loci (eQTL) analysis across multiple brain regions to examine the biological context of identified variants. Comparative analysis with conventional sliding window approaches was performed to evaluate the identification of AD-associated variants.

Application of Deep-Block to a large-scale WGS dataset from the Alzheimer’s Disease Sequencing Project (ADSP) Release 3, comprising 7,416 non-Hispanic white participants, demonstrated its capacity to effectively manage complex genomic data and identify single nucleotide polymorphisms (SNPs) as associated with AD. The Deep-Block framework identified AD-associated genetic loci, including well-known AD SNPs such as *APOE* rs429358 and rs769449 and novel single nucleotide polymorphisms (SNPs) not previously reported in AD genetic association studies, particularly within the top-performing LD blocks. The identified variants were examined through tissue-specific expression analysis to investigate their potential relationship with AD pathogenesis.

## 2 Methods

### 2.1 Data Collection and Quality Control

The ADSP participants were classified as AD cases or cognitively normal controls based on a comprehensive diagnostic framework. AD diagnoses were established through clinical diagnostic criteria, including detailed medical history, cognitive evaluations, and neuropsychological testing. Where available, these clinical assessments were supplemented with biomarker data, including cerebrospinal fluid measurements of amyloid-beta and tau proteins, and neuroimaging data indicating amyloid deposition. The ADSP participants have WGS data sequenced using multiple platforms, including IlluminaHiSeq2000 and IlluminaHiSeqXTen. This release (R3) includes 16,906 whole-genome sequences (WGS), processed and curated as part of the project. The release contains CRAMs, gVCFs, and quality-controlled project-level VCFs (pVCFs) for autosomal biallelic single nucleotide variants (SNVs) and indels, along with structural variant (SV) calls generated by Manta, Smoove, and Strelka variant callers. The WGS data were called by the Genome Center for Alzheimer’s Disease (GCAD) using VCPA 1.1, a functionally equivalent CCDG/TOPMed pipeline. WGS data underwent comprehensive quality control (QC) procedures, including SNP call rates > 95%, Hardy-Weinberg equilibrium P values < 1 × 10^-6, minor allele frequencies (MAF) > 1%, absence of sex mismatches, and sample call rates > 95%. To mitigate false associations due to population stratification, the study analyzed genome-wide genotyping data from 7,416 non-Hispanic White (NHW) participants (3,150 cognitively normal individuals (CN) and 4,266 AD patients), encompassing 10,764,329 SNPs. The male sex ratio was 56.3% for AD patients (mean age 70.1 years) and 60.7% for CN individuals (mean age 80.2 years).

### 2.2 Algorithm Implementation and Analysis

The Deep-Block framework employs a structured, three-stage process to analyze large-scale WGS data:

#### 2.2.1. Segmentation of whole genome into LD blocks

Following QC, the WGS dataset was segmented into linkage disequilibrium (LD) blocks using Plink software (v1.90b6.21 64-bit). The parameters were set as follows: the LD measure was r^2^ with a threshold of 0.9, window size of 50 variants, and maximum window physical size of 100 kilobases. LD blocks were then identified based on the genomic positions of SNPs and the extent of LD between adjacent SNPs. This configuration identified 30,218 LD Blocks, forming the basis for subsequent analyses.

##### 2.2.1.1. Comparative Analysis of Segmentation Approaches

To evaluate the effectiveness of the LD block-based segmentation approach, we conducted experiments with baseline methods using the top 1,500 LD blocks. A sliding window approach ^10^ using fixed windows of 200 variants was implemented as the baseline for comparison. For each method, SNPs were prioritized based on their computed importance scores: Deep-Block with LD-based segmentation utilized phenotype influence scores, while Random Forest and TabNet with fixed window segmentation used their respective feature importance metrics.

#### 2.2.2. Imputation of missing genotype data

Deep-Block utilizes machine learning approaches to impute missing genotype data within the LD blocks, a method supported by recent studies ^15–17^. To identify the most suitable imputer for imputing missing genotype data, preliminary experiments were conducted on the *APOE* gene region within the ADSP WGS dataset, assuming SNPs of this region are contained within LD blocks. The ADSP R3 WGS data, comprising 16,869 individuals, included 793 variants from the *APOE* gene region. After QC, the missing data proportion in this region was 4.14E-03. For the performance assessment of imputers, the missing rate was artificially increased to 8.70E-03. The modified dataset was then processed using the TopMed Imputer, establishing a benchmark for comparing the efficiency of other imputation methods. Several imputers were used: 1-NN, 5-NN, 10-NN, GAN, Iterative, MissForest ^20^, and Simple Imputer. All methods were applied to data with the same artificially increased proportion of missing genotype data to ensure consistent evaluation. The scikit-learn package ^21^ was used for machine learning imputers and the GAIN package ^22^ for the GAN Imputer.

The Simple Imputer utilizes mean, median, or mode imputation to fill missing values with the most representative statistic of the available data. The k-Nearest Neighbors (k-NN) Imputers (1-NN, 5-NN, and 10-NN) leverage data point similarity to impute missing values based on the nearest neighbors’ values. The GAN Imputer uses Generative Adversarial Networks to produce synthetic data mimicking the original data distribution, thus imputing missing values. The Iterative Imputer employs a round-robin approach, modeling each feature with missing values as a function of other features stepwise, capturing complex interactions and dependencies. The MissForest Imputer utilizes a Random Forest approach, leveraging multiple decision trees to accurately predict missing values.

The performance of these methods was evaluated using five well-established metrics: accuracy, Root Mean Squared Error (RMSE), R-squared (R2), Mean Absolute Error (MAE), and Normalized RMSE (NRMSE). The accuracy quantifies the proportion of correctly imputed values, directly reflecting an imputer’s performance. The RMSE measures the average magnitude of imputation errors, providing a straightforward accuracy metric. The R2 indicates the proportion of variance in the original data explained by the imputed data, offering insights into the imputation method’s ability to preserve data structure. The MAE calculates the average absolute error between imputed and actual values, presenting error distribution without directional bias. The NRMSE normalizes RMSE to the dataset range, facilitating the performance comparison across differently scaled datasets.

#### 2.2.3. Identification of Key LD blocks and phenotype association

The final stage identifies key LD blocks as significantly associated with the AD phenotype using TabNet, a deep learning model optimized for efficient tabular data processing ^18^. TabNet was selected for its key advantages: simultaneous feature selection and engineering capabilities, interpretability through sparse attention mechanisms, and optimized architecture for large-scale tabular data processing. Given the high-dimensional nature of our genetic data (10,764,329 variants across 30,218 LD blocks), we implemented several key strategies to prevent overfitting. These include L1 regularization (lambda_sparse=1e-3) to enforce sparsity in feature selection, learning rate scheduling with step decay (step_size=10, gamma=0.9) to optimize model convergence, and early stopping with patience monitoring to prevent unnecessary model complexity. For model validation, we maintained consistent 80/20 train-validation splits across all analyses. To validate this choice, we conducted a comparative analysis between TabNet and Random Forest approaches. The genome was segmented into 53,822 windows, each containing 200 variants, ensuring comprehensive coverage. Both models were applied to each window to assess prediction accuracy. The top 1,500 windows were selected based on accuracy metrics for each model, and feature importance was calculated for 299,822 SNPs within these windows using each model’s respective feature importance metrics. Incremental addition of top-ranked features showed that TabNet consistently outperformed Random Forest in AUC scores (average AUC of 0.56 versus 0.54), supporting our model selection. TabNet’s architecture combines the interpretability of decision tree-based models with deep learning capabilities, featuring an encoder-decoder structure, feature transformers, and attentive transformers. TabNet’s encoder processes raw tabular data, selecting relevant features through a sequential multi-step procedure using feature transformers. These transformers apply non-linear transformations to enhance the model’s learning capabilities. The attentive transformer, a key encoder component, employs the sparsemax normalization function to focus selectively on the most relevant features, optimizing model interpretability and efficiency. This stage uses TabNet to identify LD Blocks with high phenotypic relevance, focusing on features critically associated with AD. TabNet’s decoder reconstructs features from the original dataset, identifying key features within the top LD Blocks. This process assigns PIS to significant features, reflecting their phenotypic impact. The method integrates TabNet’s feature importance metrics with the Mean Decrease Impurity (MDI) metric from Random Forest, offering a systematic approach to understanding genetic influences on phenotypic traits (**Figure 1**).

**Figure 1.**
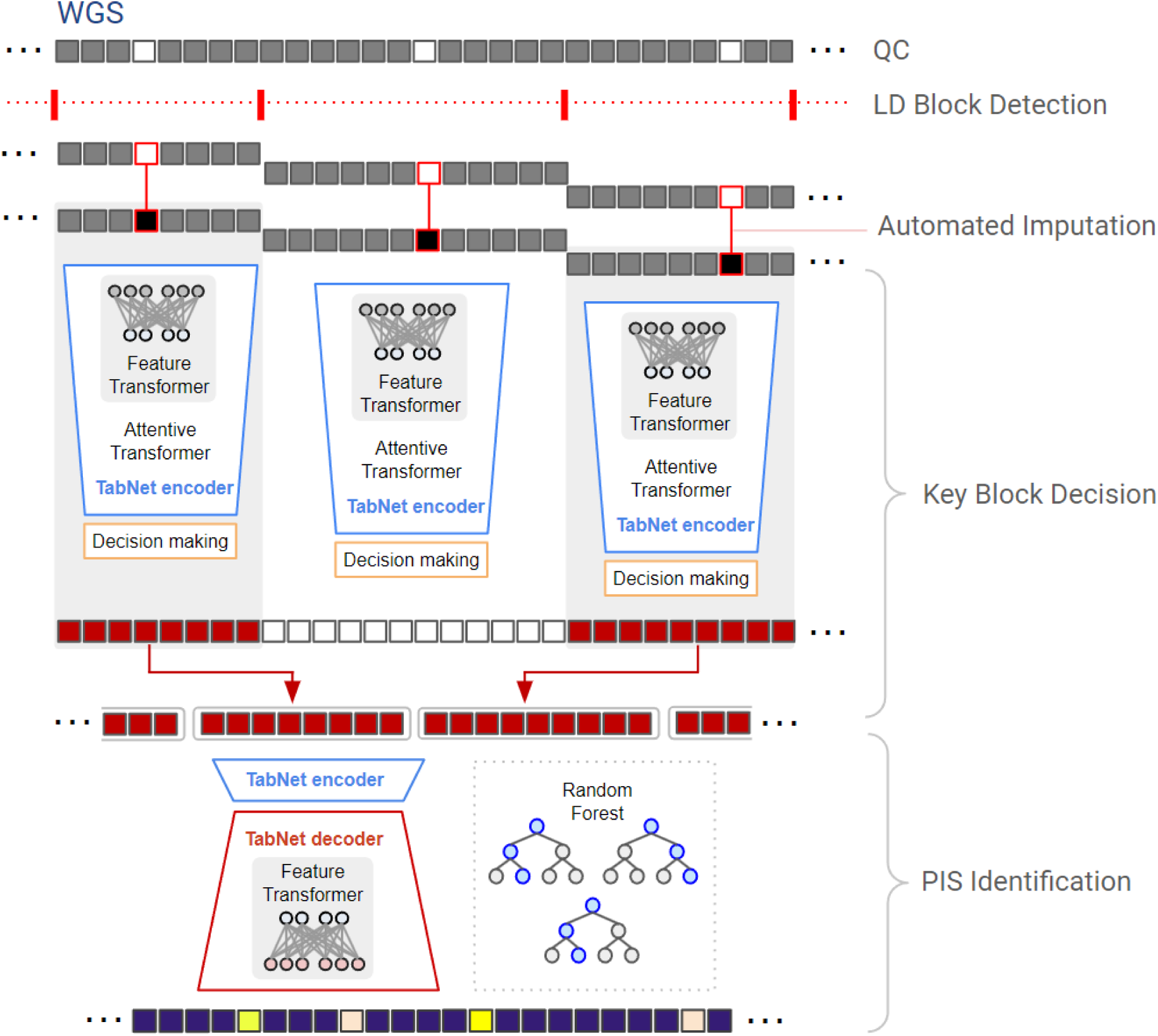
Overview of the Deep-Block Framework. This figure illustrates the sequential stages of the Deep-Block framework used in the analysis of large-scale whole genome sequencing (WGS) data for Alzheimer’s disease (AD). The process initiates with the quality control procedure (QC) of WGS data, ensuring the integrity and reliability of the genetic information. Subsequently, the data is organized into Linkage Disequilibrium (LD) blocks, indicated by red dotted lines, which reflect the partitioning based on LD parameters. The next phase, Automated imputation, is visualized as various modules corresponding to different machine learning-based imputation techniques each tasked with estimating and inputting missing genomic data. Following imputation, the TabNet encoder’s role in decision-making is depicted, using feature transformers and attentive transformers to select and prioritize LD blocks that show significant associations with AD. The final element of the diagram focuses on the identification of the Phenotype Influence Score (PIS) using the TabNet decoder in conjunction with Random Forest metrics.

This approach combines the strengths of both metrics to identify the most significant AD-associated genetic markers, offering a robust method for detecting key genetic markers within LD blocks. The PIS is calculated using the following combined formula:

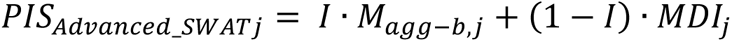

where *I* is an indicator variable that is automatically set to 1 when the TabNet model yields a higher predictive accuracy in phenotype-related classification using previously selected features, and is automatically set to 0 when the Random Forest algorithm shows superior performance in the same task. *M*_*agg-b,j*_, where b represents the batch index for the current training iteration, the feature importance from the TabNet model, represents the aggregate feature importance mask for the j-th feature. The calculation uses the total number of decision steps (N), the learning rate at each decision step (*η*_*b*_[*i*]), and a binary mask (*M*_*b*,*j*_[*i*]) that is set to 1 if the j-th feature is utilized at the i-th decision step, and 0 otherwise. Here, D represents the total number of features. The corresponding formula is as follows:

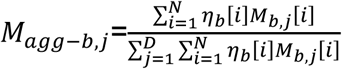

*MDI*_*j*_, the Mean Decrease Impurity from the Random Forest algorithm, quantifies the impurity reduction for a specific SNP (*SNP*_*j*_). This calculation encompasses the total number of decision trees in the model (*N*_*trees*_), each tree (t), and the node (i), using *SNP*_*j*_ for splitting, includes the number of samples at node i before the splitting 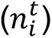 and the impurity reduction at this node 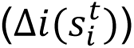. The MDI is calculated as follows:

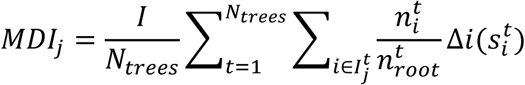

### 2.3 Functional Analysis and Annotation

We employed multiple approaches to characterize the functional implications of identified variants. The GTEx v10 database was used to identify eQTL signals across 13 brain-specific tissues, including the hypothalamus, hippocampus, cerebellum, cortex, nucleus accumbens, substantia nigra, anterior cingulate cortex, putamen, amygdala, cerebellar hemisphere, frontal cortex, spinal cord, and caudate. Statistical significance was assessed at p < 0.05. The Variant Effect Predictor (VEP) with GRCh38 human genome assembly and ANNOVAR were used to determine the genomic context of the variants.

## 3 Results

This study analyzed large-scale WGS data from the ADSP, comprising 7,416 non-Hispanic White individuals (4,266 with Alzheimer’s disease and 3,150 cognitively normal older adults). After quality control procedures, several imputation methods were comparatively evaluated: Simple, GAN, 1-NN, 5-NN, 10-NN, Iterative, MissForest, and TopMed Imputers. The assessment utilized metrics including accuracy, Root Mean Squared Error (RMSE), R-squared (R2), Mean Absolute Error (MAE), and Normalized RMSE (NRMSE).

The MissForest Imputer demonstrated superior performance among the machine learning-based methods, achieving the highest accuracy (0.999359), lowest RMSE (0.0039), and highest R2 (0.9993). The 5-NN and 10-NN Imputers also performed well, with accuracy rates of 0.999734 and 0.999626, R2 values of 0.9993, and RMSEs of 0.004 and 0.0041, respectively. The TopMed imputation server achieved an accuracy of 0.996416, RMSE of 0.0047, and R2 of 0.9081. While effective in reducing RMSE, it showed a lower capacity to capture dataset variance compared to the leading machine learning methods (**Figure 2**, **Table 1**).

**Figure 2.**
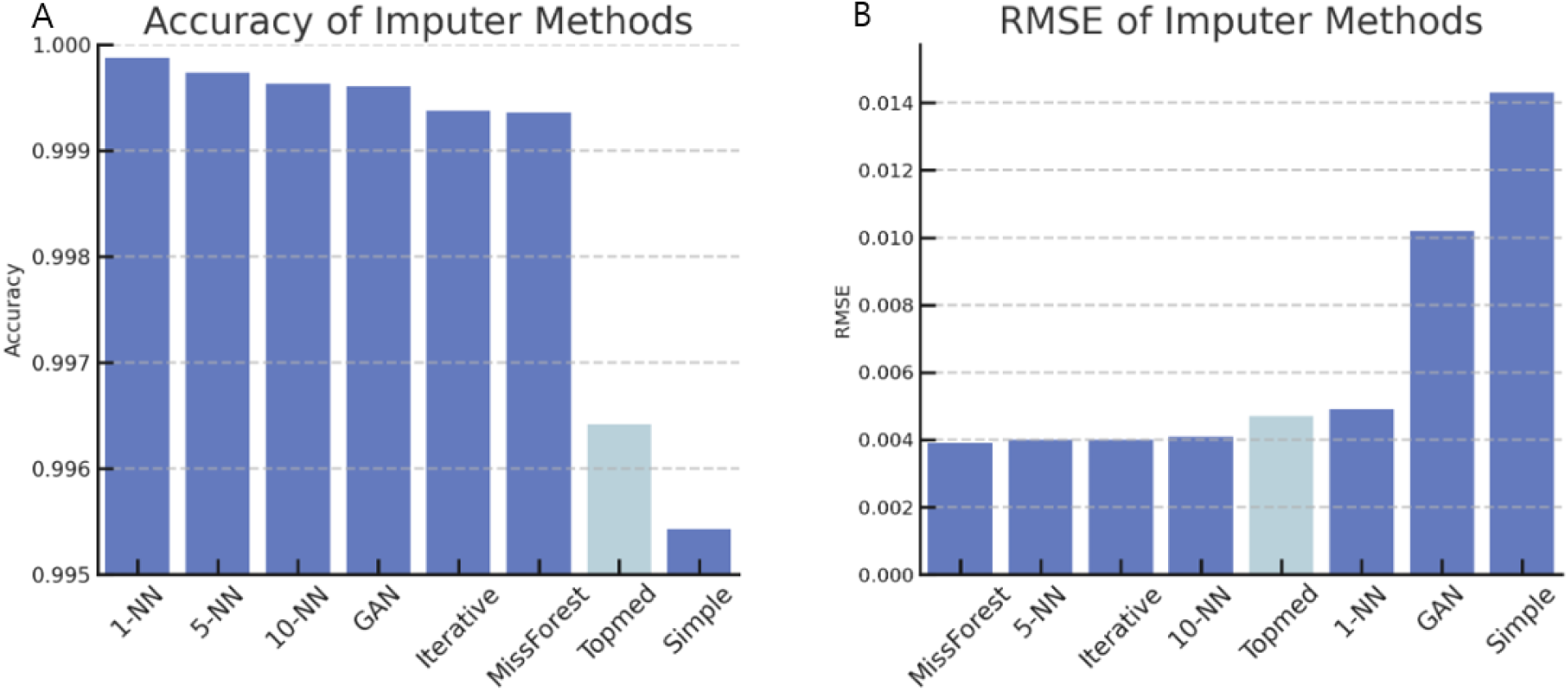
Comparative performance of imputation methods in WGS data. Fig. 2A illustrates the accuracy for several imputation methods, which reflects the proportion of correctly imputed genotypes to the total number of predictions made. A value closer to 1 denotes a higher rate of correct imputations. In this analysis, the 1-NN Imputer exhibits the highest accuracy, while the Simple Imputer shows the least accuracy, pointing to a greater discrepancy in its predictions. Fig. 2B displays the Root Mean Square Error (RMSE) across the imputation methods, a metric for quantifying the average errors in the predicted values. The lower the RMSE, the more accurate the imputation. Here, the MissForest Imputer emerges as the most accurate with the smallest RMSE, while the Simple Imputer displays the largest RMSE, indicative of lower accuracy. The results of the Topmed Imputer were not as pronounced, falling behind with lower accuracy and a higher RMSE than several other imputers.

**Table 1.** Comparison of imputation efficacies of imputation methods. The table shows performance metrics for several imputation methods of missing genotypes. The accuracy measures the proportion of correctly imputed genotypes, where the 1-NN Imputer ranks the highest, suggesting the greatest precision in imputation among the methods. The Root Mean Squared Error (RMSE) shows the MissForest Imputer as the most accurate, with the smallest values indicating minimal deviation from actual data. R-squared (R2) values for the 5-NN, 10-NN, Iterative, and MissForest Imputers indicate that these models account for a significant portion of the variance, suggesting a strong correlation with the observed data. The Mean Absolute Error (MAE) is lowest for the 1-NN, 5-NN, and MissForest Imputers, indicating higher precision. The Normalized Root Mean Squared Error (NRMSE) further confirms the MissForest Imputer’s superior performance. Overall, the MissForest Imputer exhibits the highest precision in imputation of missing genotypes.

| IMPUTER METHOD | ACCURACY | RMSE | R2 | MAE | NRMSE |
| --- | --- | --- | --- | --- | --- |
| 1-NN IMPUTER | 0.999875 | 0.0049 | 0.9989 | 0 | 0.0049 |
| 5-NN IMPUTER | 0.999734 | 0.004 | 0.9993 | 0 | 0.004 |
| 10-NN IMPUTER | 0.999626 | 0.0041 | 0.9993 | 0.0001 | 0.0041 |
| GAN IMPUTER | 0.999606 | 0.0102 | 0.9981 | 0.0002 | 0.0102 |
| ITERATIVE IMPUTER | 0.999373 | 0.004 | 0.9993 | 0.0001 | 0.004 |
| MISSFOREST IMPUTER | 0.999359 | 0.0039 | 0.9993 | 0 | 0.0039 |
| TOPMED IMPUTER | 0.996416 | 0.0047 | 0.9081 | 0.0002 | 0.0047 |
| SIMPLE IMPUTER | 0.995432 | 0.0143 | 0.9965 | 0.0006 | 0.0143 |

Computation time for imputation methods was crucial due to the large-scale WGS data. **Table S1** shows that the MissForest Imputer required up to 327 seconds for the largest block size, significantly longer than the 5-NN Imputer, which processed the same block in just over 50 milliseconds. Balancing imputation accuracy and processing efficiency, the 5-NN Imputer was selected as the most suitable imputation method for our dataset. This choice was based on its high accuracy, and fast LD blocks were determined using Plink, resulting in 30,218 LD blocks with an average size of 388 genetic variants.

Genomic regions not covered by LD blocks, comprising only 0.19% of the genome, were excluded from the analysis due to their negligible size. **Figure 3B** demonstrates the correlation between the number of blocks per chromosome and chromosome length. TabNet was then applied to the LD blocks to assess phenotype prediction accuracy using a binary classification model. Blocks were ranked based on prediction accuracy, and as the number of blocks increased, the analysis showed that the number of important features with non-zero TabNet feature importance did not increase significantly. **Figure 3A** illustrates this by depicting genetic variants within the key blocks as blue bars. The analysis extended up to 1500 blocks, revealing a plateau in the count of important features, indicating that the critical variants for phenotype prediction were already captured within the initial top blocks. This suggests that further analysis beyond these blocks may not provide additional meaningful insights.

**Figure 3.**
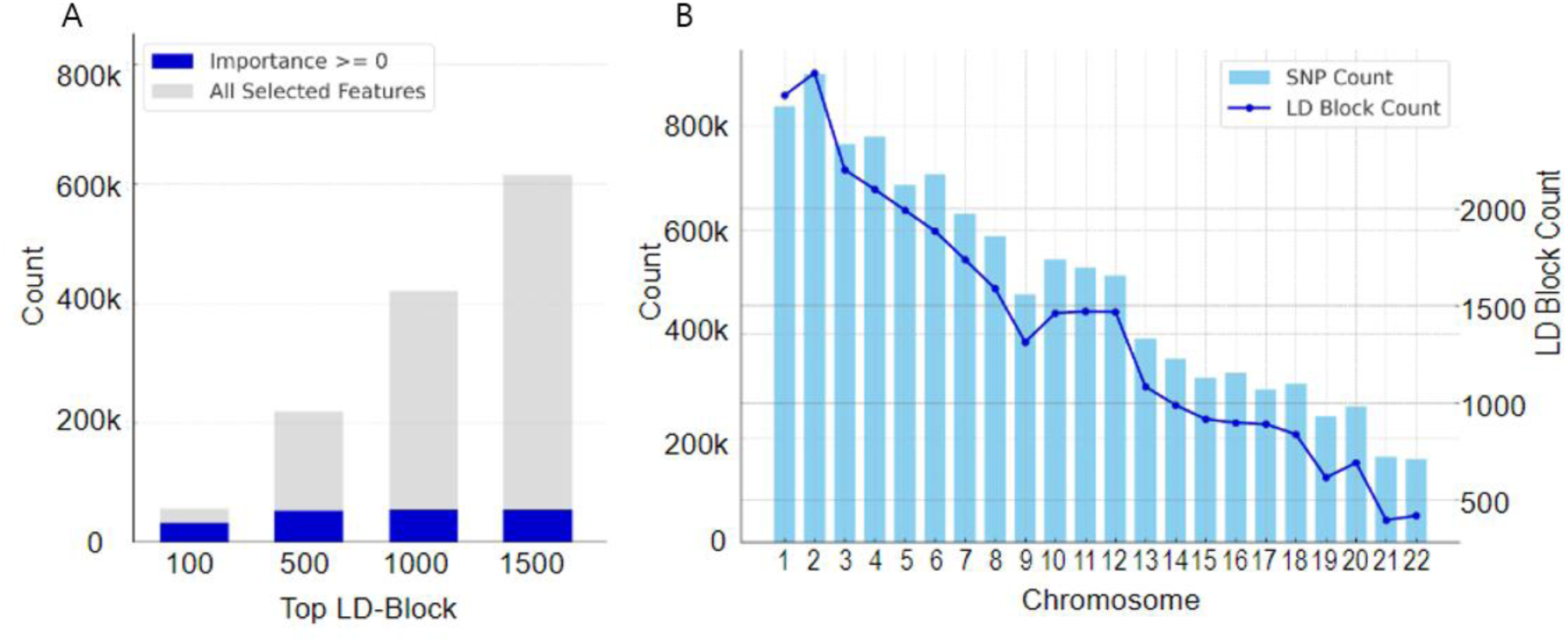
Feature importance and distribution across LD blocks in the ADSP WGS Dataset. In Fig. 3A, the analysis determined the feature importance using TabNet for the top 100 to 1500 LD blocks. The blue bars represent genetic variants within these blocks, where TabNet was assigned a feature importance greater than zero, indicating their relevance in phenotype prediction. The gray bars indicate all selected features, regardless of their importance score. The steady count of important features across increasing block ranks suggests that the most critical variants for phenotype prediction were concentrated in the top blocks. Fig 3B visualizes the distribution of LD blocks and SNP counts across chromosomes. The bar chart demonstrates that the number of LD blocks and SNPs is proportionate to the chromosome length, with larger chromosomes containing more blocks.

To evaluate our LD block-based approach, we conducted a comparison with baseline segmentation methods using the top 1,500 LD blocks. For each method, we incrementally selected SNPs based on their importance scores and assessed their predictive performance. Analysis showed that the Deep-Block framework achieved an average AUC of 0.66, while fixed sliding window-based approaches using Random Forest and TabNet showed average AUCs of 0.54 and 0.56, respectively. This pattern was consistent across different numbers of selected SNPs, as visualized in **Figure 4A**, which presents the AUC scores according to the number of SNPs used in the analysis for each method.

**Figure 4.**
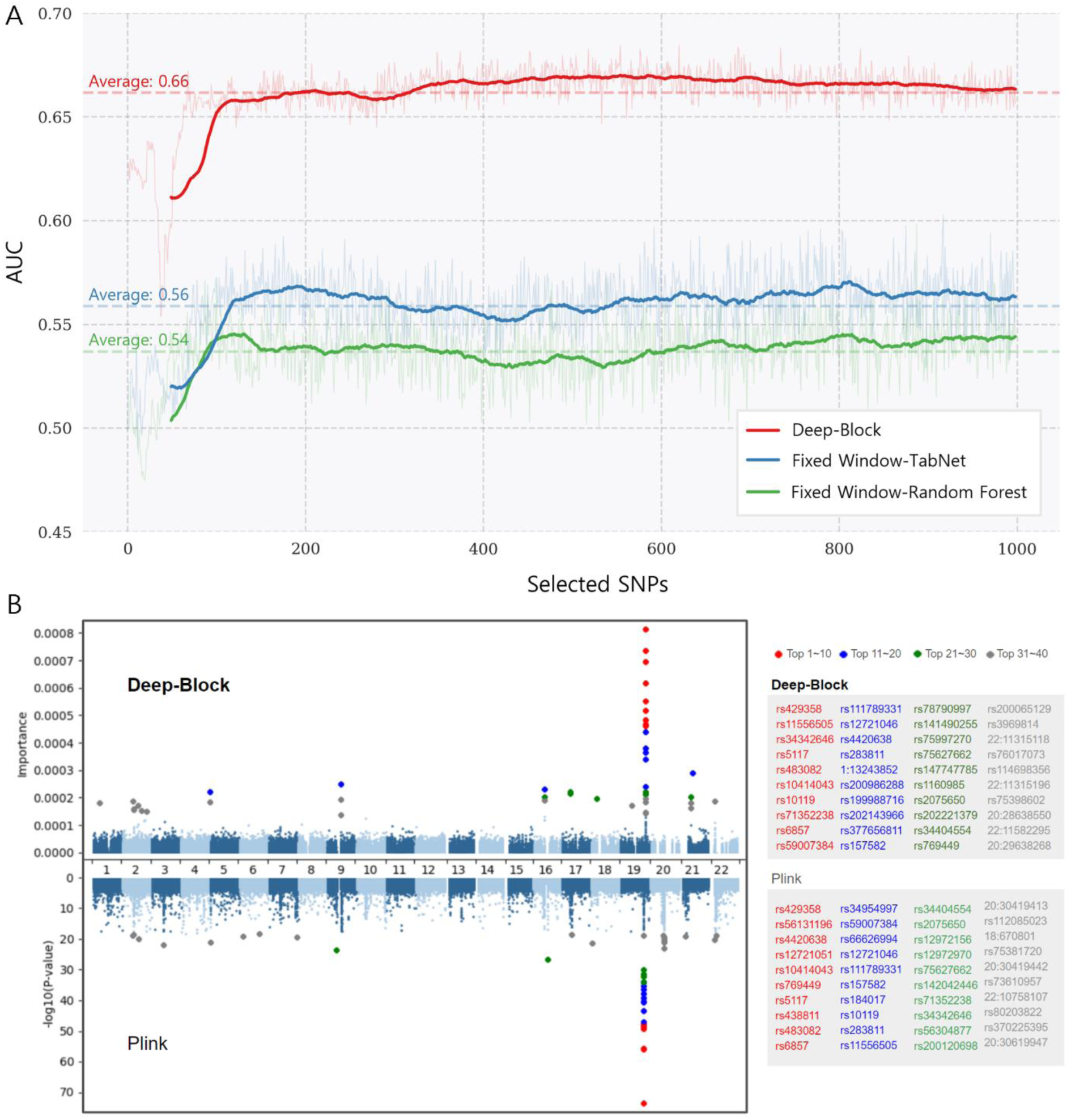
Performance and genomic analysis of Deep-Block framework. (A) Performance comparison between different genomic segmentation approaches, showing AUC scores based on selected SNPs from the top 1,500 LD blocks. Lines represent smoothed AUC scores for Deep-Block with LD-based segmentation (red), TabNet (blue) and Random Forest (green) with fixed window approaches. (B) Miami plot comparing genetic association statistics between Deep-Block (upper) and Plink (lower). The plots display genome-wide SNP significance, with chromosomes listed numerically on the horizontal axis. Color-coding indicates significance ranking: red (top 1-10), blue (11-20), green (21-30), and gray (31-40), with a side panel listing SNPs in each category.

The study analyzed 615,282 genetic variants within the top 1500 blocks, calculating the PIS for each genetic variant using the methodology outlined in the Methods section. **Table 2** presents the top 30 genetic variants with the highest PIS. The SNP rs429358 within the *APOE* gene on chromosome 19, a well-known AD risk SNP ^23^, demonstrated the highest importance score. Other high-importance SNPs on chromosome 19 include rs11556505 in *TOMM40* ^24^) and rs34342646 in *NECTIN2* ^25^, further emphasizing the relevance of chromosome 19 in AD. The study also confirmed previously reported AD-associated SNPs, including rs5117 and rs483082 in *APOC1*, and rs10414043 (*APOC1*), rs10119 (*TOMM40*), and rs71352238 (*TOMM40*).

**Table 2.** SNPs with highest phenotype influence scores (PIS) associated with AD. This table catalogs the top 30 single nucleotide polymorphisms (SNPs) ranked by PIS, derived from an extensive examination of 54,949 genetic variants within the top 1500 LD blocks using TabNet. The highest-scoring SNPs are predominantly located on chromosome 19, related to genes such as APOE, APOC1, NECTIN2, and TOMM40—well-established AD-associated genes. Additionally, this table includes novel findings, highlighting SNPs and genes previously unidentified in AD genetic association studies.

| RANK | FEATURE | CHR. | POSITION | VARIATION | GENE | IMPORTANCE | REFERENCE |
| --- | --- | --- | --- | --- | --- | --- | --- |
| 1 | rs429358 | 19 | 44908683 | T/C | APOE | 8.00E-04 | 22 |
| 2 | rs11556505 | 19 | 44892886 | C/T | TOMM40 | 7.23E-04 | 24 |
| 3 | rs34342646 | 19 | 44884872 | G/A | NECTIN2 | 6.85E-04 | 25 |
| 4 | rs5117 | 19 | 44915532 | T/C | APOC1 | 6.85E-04 |  |
| 5 | rs483082 | 19 | 44912920 | G/T | APOC1 | 6.09E-04 | 34 |
| 6 | rs10414043 | 19 | 44912455 | G/A | APOC1 | 5.43E-04 | 35 |
| 7 | rs10119 | 19 | 44903415 | G/A | TOMM40 | 5.11E-04 | 24 |
| 8 | rs71352238 | 19 | 44891078 | T/C | TOMM40 | 4.76E-04 | 35 |
| 9 | rs6857 | 19 | 44888996 | C/T | NECTIN2 | 4.63E-04 | 24 |
| 10 | rs59007384 | 19 | 44893407 | G/A G/T | TOMM40 | 4.57E-04 | 35 |
| 11 | rs111789331 | 19 | 44923867 | T/A |  | 4.34E-04 |  |
| 12 | rs12721046 | 19 | 44917996 | G/A | APOC1 | 3.75E-04 | 36 |
| 13 | rs4420638 | 19 | 44919688 | A/G | APOC1 | 3.61E-04 | 37 |
| 14 | rs283811 | 19 | 44885242 | A/C A/G | NECTIN2 | 3.37E-04 | 25 |
| 15 | 1:13243852 | 1 | 13243852 | C/T |  | 3.06E-04 |  |
| 16 | rs200986288 | 20 | 30185632 | A/C A/T |  | 2.87E-04 |  |
| 17 | rs199988716 | 2 | 95935492 | G/A |  | 2.51E-04 |  |
| 18 | rs202143966 | 9 | 63769614 | T/C |  | 2.49E-04 |  |
| 19 | rs377656811 | 22 | 16427107 | C/A C/T |  | 2.45E-04 |  |
| 20 | rs157582 | 19 | 44892961 | C/T | TOMM40 | 2.37E-04 | 38 |
| 21 | rs78790997 | 16 | 33736499 | C/G | LOC107984083 | 2.28E-04 |  |
| 22 | rs141490255 | 1 | 58630472 | G/A |  | 2.27E-04 |  |
| 23 | rs75997270 | 4 | 189924739 | C/A C/G | LOC728339 | 2.21E-04 |  |
| 24 | rs75627662 | 19 | 44910318 | C/T |  | 2.20E-04 |  |
| 25 | rs147747785 | 17 | 22046134 | G/T |  | 2.20E-04 |  |
| 26 | rs1160985 | 19 | 44900154 | C/T | TOMM40 | 2.18E-04 | 39 |
| 27 | rs2075650 | 19 | 44892361 | A/G | TOMM40 | 2.16E-04 | 24 |
| 28 | rs202221379 | 17 | 22046133 | G/T |  | 2.14E-04 |  |
| 29 | rs34404554 | 19 | 44892651 | C/G | TOMM40 | 2.09E-04 | 35 |
| 30 | rs769449 | 19 | 44906744 | G/A | APOE | 2.09E-04 | 40 |

In addition to confirming the importance of known genes, this study identified novel high-importance SNPs not previously identified in genetic association studies for AD, particularly within the top 30 variants. Notably, the study identified novel variants such as rs200986288, rs199988716, and rs202143966. Additionally, previously unreported genes associated with AD were identified, including *LOC107984083* (rs78790997) and *LOC728339* (rs75997270).

**Figure 4B** displays vertically aligned, symmetrical Manhattan plots (Miami plot) from two genetic association analysis methods: Deep-Block and Plink. The upper plot depicts the PIS derived from Deep-Block, while the lower plot shows the statistical significance levels (-log10 P-value) obtained using Plink across all chromosomes. Both plots arrange chromosomes along the x-axis, offering a chromosomal position view of the analyzed genomic variants. Each plot highlights the top 40 genetic variants using color-coded dots: red for the top 1-10 genetic variants, blue for the top 11-20 genetic variants, green for the top 21-30 genetic variants, and gray for the top 31-40 genetic variants. Of note, the Deep-Block approach identified the same genetic variants that the Plink identified as well as novel genetic variants that the Plink could not identify.

To contextualize the Deep-Block findings, the identified loci were compared to established AD-linked genetic loci reported in the European Alzheimer’s and Dementia Biobank (EADB) and the recently updated GWAS catalog ^26^. The validation process benefited from ancestral homogeneity, as both our study cohort (non-Hispanic white participants) and the comparative databases are predominantly of European descent, ensuring methodological consistency in the assessment of our framework’s performance. The analysis aligned with 15 genetic loci from the GWAS catalog, distributed across various tiers (3 in Tier1, 1 in Tier2, 2 in Tier3, 2 unverified, and 3 in the ’Other’ category), as detailed in Table S1. This comparison excluded the well-studied *APOE*, *TOMM40*/*APOC1*/*NECTIN2* loci to focus on other significant genetic associations. The results were also compared against databases referenced in the GWAS catalog, including IGAP2 ^27^, PGC1 ^28^, IGAP2+UKB ^29^, GR@ACE ^30^, PGC2 ^31^, and EADB ^32^. **Table S2** highlights Tier1 genes *ABCA7*, *BIN1*, and *CR1*, identified in multiple studies. This confirms the Deep-Block method’s relevance to known AD genetic markers and indicates both confirmatory and potentially novel genetic associations with the disease.

## 4 Discussion

This study developed and applied the Deep-Block framework to large-scale WGS data from the ADSP to investigate the genetic basis of AD. The approach centered on utilizing LD blocks combined with automated imputation to improve the accuracy of genetic analysis. The PIS-based variant prioritization identified known AD-associated variants, with PIS values of 8.00E-04, 7.23E-04, and 6.85E-04 observed for established variants in *APOE* (rs429358), *TOMM40* (rs11556505), and *NECTIN2* (rs34342646) genes, respectively (Table 2). To facilitate comprehensive analysis of genetic associations and reproducibility of our findings, we have provided the complete list of 615,281 SNPs and their corresponding PIS values, along with the source code and example datasets in our Deep-Block GitHub repository (https://github.com/taehojo/Deep-Block/). The predictive performance analysis (**Figure 4A**) demonstrates that SNPs selected based on PIS values show increasing AUC scores, validating the utility of PIS rankings in variant identification. While traditional GWAS approaches rely on established p-value thresholds, our Deep-Block framework takes a more flexible approach, using known AD-associated variants as empirical benchmarks for significance assessment.

The eQTL analysis across brain regions revealed region-specific expression patterns. Our integrative approach showed that several variants had strong functional evidence. In particular, rs200986288 demonstrated widespread effects across brain regions (506 associations, average β=0.42), while rs75997270 showed consistent upregulation (P=2.75e-40, β=0.68) across analyzed brain regions. These findings suggest potential functional roles beyond mere LD-based associations, though further functional analysis through fine-mapping and cross-population studies will be valuable for confirming the functionality. The cerebellar hemisphere showed 145 eQTL associations, cerebellum 131, and nucleus accumbens basal ganglia 124. Rs75997270, an intronic variant in *FRG1-DT* (lncRNA), had a significant association (P=2.75e-40) in the nucleus accumbens and was detected across all analyzed brain regions. The *APOE* downstream variant rs75627662 and *APOC1* intronic variant rs5117 were detected in the nucleus accumbens basal ganglia. Rs200986288 was detected in 506 eQTL associations across the analyzed brain regions. The VEP and ANNOVAR analyses showed rs75627662 as an *APOE* downstream variant, rs5117 as an *APOC1* intronic variant, rs111789331 as an upstream variant of *APOC1P1*, and rs75997270 as an intronic variant within *FRG1-DT*.

The analysis also identified additional genetic variants and genes (e.g., rs199988716, *LOC107984083*, and *ANKRD30BL*) associated with AD. However, this study used sequencing data from non-Hispanic white individuals, which may limit the broad applicability of the findings. As more ancestrally diverse cohorts become available in the ADSP, we look forward to extending these analyses to ensure broader relevance and applicability of findings across different populations.

### Conclusion

This study developed and applied the Deep-Block AI framework to large-scale ADSP WGS data for genetic association analysis for AD. The approach involved segmenting the whole genome into LD blocks and applying automated imputation of missing genotypes for data preprocessing. The Deep-Block framework identified AD-associated genetic loci, including both previously identified and novel SNPs, supported by tissue-specific eQTL evidence across brain regions. The framework integrated eQTL analysis across brain regions and was compared with sliding window approaches in variant identification. Compared to traditional methods such as Plink and SKAT-O ^33^, Deep-Block uses LD structure and attention-based feature selection to analyze high-dimensional genomic data more comprehensively, potentially capturing genetic interactions that are not detected by conventional approaches. Unlike SWAT-CNN ^10^, which utilizes fixed genomic fragments, Deep-Block segments the genome based on LD-defined regions, thereby improving the identification of biologically relevant patterns. This framework demonstrates its capability to analyze large-scale genomic data effectively and identified both known and novel genetic variants associated with Alzheimer’s disease.

## Data Availability

All data produced in the present study are available upon reasonable request to the authors.

## Funding

This research was partially supported by the Alzheimer’s Association (AA) under the grant number AARG 22-974053 and the National Institutes of Health (NIH): P30 AG010133, P30 AG072976, R01 AG019771, R01 AG057739, U01 AG024904, R01 LM013463, R01 AG068193, T32 AG071444, U01 AG068057, U01AG072177, U19AG074879, R03AG063250, and R01 LM012535.

The Alzheimer’s Disease Sequencing Project (ADSP) is comprised of two Alzheimer’s Disease (AD) genetics consortia and three National Human Genome Research Institute (NHGRI) funded Large Scale Sequencing and Analysis Centers (LSAC). The two AD genetics consortia are the Alzheimer’s Disease Genetics Consortium (ADGC) funded by NIA (U01 AG032984), and the Cohorts for Heart and Aging Research in Genomic Epidemiology (CHARGE) funded by NIA (R01 AG033193), the National Heart, Lung, and Blood Institute (NHLBI), other National Institute of Health (NIH) institutes and other foreign governmental and non-governmental organizations. The Discovery Phase analysis of sequence data is supported through UF1AG047133 (to Drs. Schellenberg, Farrer, Pericak-Vance, Mayeux, and Haines); U01AG049505 to Dr. Seshadri; U01AG049506 to Dr. Boerwinkle; U01AG049507 to Dr. Wijsman; and U01AG049508 to Dr. Goate and the Discovery Extension Phase analysis is supported through U01AG052411 to Dr. Goate, U01AG052410 to Dr. Pericak-Vance and U01 AG052409 to Drs. Seshadri and Fornage.

Sequencing for the Follow Up Study (FUS) is supported through U01AG057659 (to Drs. PericakVance, Mayeux, and Vardarajan) and U01AG062943 (to Drs. Pericak-Vance and Mayeux). Data generation and harmonization in the Follow-up Phase is supported by U54AG052427 (to Drs. Schellenberg and Wang). The FUS Phase analysis of sequence data is supported through U01AG058589 (to Drs. Destefano, Boerwinkle, De Jager, Fornage, Seshadri, and Wijsman), U01AG058654 (to Drs. Haines, Bush, Farrer, Martin, and Pericak-Vance), U01AG058635 (to Dr. Goate), RF1AG058066 (to Drs. Haines, Pericak-Vance, and Scott), RF1AG057519 (to Drs. Farrer and Jun), R01AG048927 (to Dr. Farrer), and RF1AG054074 (to Drs. Pericak-Vance and Beecham).

The ADGC cohorts include: Adult Changes in Thought (ACT) (U01 AG006781, U19 AG066567), the Alzheimer’s Disease Research Centers (ADRC) (P30 AG062429, P30 AG066468, P30 AG062421, P30 AG066509, P30 AG066514, P30 AG066530, P30 AG066507, P30 AG066444, P30 AG066518, P30 AG066512, P30 AG066462, P30 AG072979, P30 AG072972, P30 AG072976, P30 AG072975, P30 AG072978, P30 AG072977, P30 AG066519, P30 AG062677, P30 AG079280, P30 AG062422, P30 AG066511, P30 AG072946, P30 AG062715, P30 AG072973, P30 AG066506, P30 AG066508, P30 AG066515, P30 AG072947, P30 AG072931, P30 AG066546, P20 AG068024, P20 AG068053, P20 AG068077, P20 AG068082, P30 AG072958, P30 AG072959), the Chicago Health and Aging Project (CHAP) (R01 AG11101, RC4 AG039085, K23 AG030944), Indiana Memory and Aging Study (IMAS) (R01 AG019771), Indianapolis Ibadan (R01 AG009956, P30 AG010133), the Memory and Aging Project (MAP) (R01 AG17917), Mayo Clinic (MAYO) (R01 AG032990, U01 AG046139, R01 NS080820, RF1 AG051504, P50 AG016574), Mayo Parkinson’s Disease controls (NS039764, NS071674, 5RC2HG005605), University of Miami (R01 AG027944, R01 AG028786, R01 AG019085, IIRG09133827, A2011048), the Multi-Institutional Research in Alzheimer’s Genetic Epidemiology Study (MIRAGE) (R01 AG09029, R01 AG025259), the National Centralized Repository for Alzheimer’s Disease and Related Dementias (NCRAD) (U24 AG021886), the National Institute on Aging Late Onset Alzheimer’s Disease Family Study (NIA-LOAD) (U24 AG056270), the Religious Orders Study (ROS) (P30 AG10161, R01 AG15819), the Texas Alzheimer’s Research and Care Consortium (TARCC) (funded by the Darrell K Royal Texas Alzheimer’s Initiative), Vanderbilt University/Case Western Reserve University (VAN/CWRU) (R01 AG019757, R01 AG021547, R01 AG027944, R01 AG028786, P01 NS026630, and Alzheimer’s Association), the Washington Heights-Inwood Columbia Aging Project (WHICAP) (RF1 AG054023), the University of Washington Families (VA Research Merit Grant, NIA: P50AG005136, R01AG041797, NINDS: R01NS069719), the Columbia University Hispanic Estudio Familiar de Influencia Genetica de Alzheimer (EFIGA) (RF1 AG015473), the University of Toronto (UT) (funded by Wellcome Trust, Medical Research Council, Canadian Institutes of Health Research), and Genetic Differences (GD) (R01 AG007584). The CHARGE cohorts are supported in part by National Heart, Lung, and Blood Institute (NHLBI) infrastructure grant HL105756 (Psaty), RC2HL102419 (Boerwinkle) and the neurology working group is supported by the National Institute on Aging (NIA) R01 grant AG033193.

The CHARGE cohorts participating in the ADSP include the following: Austrian Stroke Prevention Study (ASPS), ASPS-Family study, and the Prospective Dementia Registry-Austria (ASPS/PRODEM-Aus), the Atherosclerosis Risk in Communities (ARIC) Study, the Cardiovascular Health Study (CHS), the Erasmus Rucphen Family Study (ERF), the Framingham Heart Study (FHS), and the Rotterdam Study (RS). ASPS is funded by the Austrian Science Fond (FWF) grant number P20545-P05 and P13180 and the Medical University of Graz. The ASPS-Fam is funded by the Austrian Science Fund (FWF) project I904), the EU Joint Programme – Neurodegenerative Disease Research (JPND) in frame of the BRIDGET project (Austria, Ministry of Science) and the Medical University of Graz and the Steiermärkische Krankenanstalten Gesellschaft. PRODEM-Austria is supported by the Austrian Research Promotion agency (FFG) (Project No. 827462) and by the Austrian National Bank (Anniversary Fund, project 15435. ARIC research is carried out as a collaborative study supported by NHLBI contracts (HHSN268201100005C, HHSN268201100006C, HHSN268201100007C, HHSN268201100008C, HHSN268201100009C, HHSN268201100010C, HHSN268201100011C, and HHSN268201100012C). Neurocognitive data in ARIC is collected by U01 2U01HL096812, 2U01HL096814, 2U01HL096899, 2U01HL096902, 2U01HL096917 from the NIH (NHLBI, NINDS, NIA and NIDCD), and with previous brain MRI examinations funded by R01-HL70825 from the NHLBI. CHS research was supported by contracts HHSN268201200036C, HHSN268200800007C, N01HC55222, N01HC85079, N01HC85080, N01HC85081, N01HC85082, N01HC85083, N01HC85086, and grants U01HL080295 and U01HL130114 from the NHLBI with additional contribution from the National Institute of Neurological Disorders and Stroke (NINDS). Additional support was provided by R01AG023629, R01AG15928, and R01AG20098 from the NIA. FHS research is supported by NHLBI contracts N01-HC-25195 and HHSN268201500001I. This study was also supported by additional grants from the NIA (R01s AG054076, AG049607 and AG033040 and NINDS (R01 NS017950). The ERF study as a part of EUROSPAN (European Special Populations Research Network) was supported by European Commission FP6 STRP grant number 018947 (LSHG-CT-2006-01947) and also received funding from the European Community’s Seventh Framework Programme (FP7/2007-2013)/grant agreement HEALTH-F4-2007-201413 by the European Commission under the programme “Quality of Life and Management of the Living Resources” of 5th Framework Programme (no. QLG2-CT-2002-01254). High-throughput analysis of the ERF data was supported by a joint grant from the Netherlands Organization for Scientific Research and the Russian Foundation for Basic Research (NWO-RFBR 047.017.043). The Rotterdam Study is funded by Erasmus Medical Center and Erasmus University, Rotterdam, the Netherlands Organization for Health Research and Development (ZonMw), the Research Institute for Diseases in the Elderly (RIDE), the Ministry of Education, Culture and Science, the Ministry for Health, Welfare and Sports, the European Commission (DG XII), and the municipality of Rotterdam. Genetic data sets are also supported by the Netherlands Organization of Scientific Research NWO Investments (175.010.2005.011, 911-03-012), the Genetic Laboratory of the Department of Internal Medicine, Erasmus MC, the Research Institute for Diseases in the Elderly (014-93-015; RIDE2), and the Netherlands Genomics Initiative (NGI)/Netherlands Organization for Scientific Research (NWO) Netherlands Consortium for Healthy Aging (NCHA), project 050-060-810. All studies are grateful to their participants, faculty and staff. The content of these manuscripts is solely the responsibility of the authors and does not necessarily represent the official views of the National Institutes of Health or the U.S. Department of Health and Human Services.

The FUS cohorts include: the Alzheimer’s Disease Research Centers (ADRC) (P30 AG062429, P30 AG066468, P30 AG062421, P30 AG066509, P30 AG066514, P30 AG066530, P30 AG066507, P30 AG066444, P30 AG066518, P30 AG066512, P30 AG066462, P30 AG072979, P30 AG072972, P30 AG072976, P30 AG072975, P30 AG072978, P30 AG072977, P30 AG066519, P30 AG062677, P30 AG079280, P30 AG062422, P30 AG066511, P30 AG072946, P30 AG062715, P30 AG072973, P30 AG066506, P30 AG066508, P30 AG066515, P30 AG072947, P30 AG072931, P30 AG066546, P20 AG068024, P20 AG068053, P20 AG068077, P20 AG068082, P30 AG072958, P30 AG072959), Alzheimer’s Disease Neuroimaging Initiative (ADNI) (U19AG024904), Amish Protective Variant Study (RF1AG058066), Cache County Study (R01AG11380, R01AG031272, R01AG21136, RF1AG054052), Case Western Reserve University Brain Bank (CWRUBB) (P50AG008012), Case Western Reserve University Rapid Decline (CWRURD) (RF1AG058267, NU38CK000480), CubanAmerican Alzheimer’s Disease Initiative (CuAADI) (3U01AG052410), Estudio Familiar de Influencia Genetica en Alzheimer (EFIGA) (5R37AG015473, RF1AG015473, R56AG051876), Genetic and Environmental Risk Factors for Alzheimer Disease Among African Americans Study (GenerAAtions) (2R01AG09029, R01AG025259, 2R01AG048927), Gwangju Alzheimer and Related Dementias Study (GARD) (U01AG062602), Hillblom Aging Network (2014-A-004-NET, R01AG032289, R01AG048234), Hussman Institute for Human Genomics Brain Bank (HIHGBB) (R01AG027944, Alzheimer’s Association “Identification of Rare Variants in Alzheimer Disease”), Ibadan Study of Aging (IBADAN) (5R01AG009956), Longevity Genes Project (LGP) and LonGenity (R01AG042188, R01AG044829, R01AG046949, R01AG057909, R01AG061155, P30AG038072), Mexican Health and Aging Study (MHAS) (R01AG018016), Multi-Institutional Research in Alzheimer’s Genetic Epidemiology (MIRAGE) (2R01AG09029, R01AG025259, 2R01AG048927), Northern Manhattan Study (NOMAS) (R01NS29993), Peru Alzheimer’s Disease Initiative (PeADI) (RF1AG054074), Puerto Rican 1066 (PR1066) (Wellcome Trust (GR066133/GR080002), European Research Council (340755)), Puerto Rican Alzheimer Disease Initiative (PRADI) (RF1AG054074), Reasons for Geographic and Racial Differences in Stroke (REGARDS) (U01NS041588), Research in African American Alzheimer Disease Initiative (REAAADI) (U01AG052410), the Religious Orders Study (ROS) (P30 AG10161, P30 AG72975, R01 AG15819, R01 AG42210), the RUSH Memory and Aging Project (MAP) (R01 AG017917, R01 AG42210Stanford Extreme Phenotypes in AD (R01AG060747), University of Miami Brain Endowment Bank (MBB), University of Miami/Case Western/North Carolina A&T African American (UM/CASE/NCAT) (U01AG052410, R01AG028786), and Wisconsin Registry for Alzheimer’s Prevention (WRAP) (R01AG027161 and R01AG054047).

The four LSACs are: the Human Genome Sequencing Center at the Baylor College of Medicine (U54 HG003273), the Broad Institute Genome Center (U54HG003067), The American Genome Center at the Uniformed Services University of the Health Sciences (U01AG057659), and the Washington University Genome Institute (U54HG003079). Genotyping and sequencing for the ADSP FUS is also conducted at John P. Hussman Institute for Human Genomics (HIHG) Center for Genome Technology (CGT).

Biological samples and associated phenotypic data used in primary data analyses were stored at Study Investigators institutions, and at the National Centralized Repository for Alzheimer’s Disease and Related Dementias (NCRAD, U24AG021886) at Indiana University funded by NIA. Associated Phenotypic Data used in primary and secondary data analyses were provided by Study Investigators, the NIA funded Alzheimer’s Disease Centers (ADCs), and the National Alzheimer’s Coordinating Center (NACC, U24AG072122) and the National Institute on Aging Genetics of Alzheimer’s Disease Data Storage Site (NIAGADS, U24AG041689) at the University of Pennsylvania, funded by NIA. Harmonized phenotypes were provided by the ADSP Phenotype Harmonization Consortium (ADSP-PHC), funded by NIA (U24 AG074855, U01 AG068057 and R01 AG059716) and Ultrascale Machine Learning to Empower Discovery in Alzheimer’s Disease Biobanks (AI4AD, U01 AG068057). This research was supported in part by the Intramural Research Program of the National Institutes of health, National Library of Medicine. Contributors to the Genetic Analysis Data included Study Investigators on projects that were individually funded by NIA, and other NIH institutes, and by private U.S. organizations, or foreign governmental or nongovernmental organizations.

The ADSP Phenotype Harmonization Consortium (ADSP-PHC) is funded by NIA (U24 AG074855, U01 AG068057 and R01 AG059716). The harmonized cohorts within the ADSP-PHC include: the Anti-Amyloid Treatment in Asymptomatic Alzheimer’s study (A4 Study), a secondary prevention trial in preclinical Alzheimer’s disease, aiming to slow cognitive decline associated with brain amyloid accumulation in clinically normal older individuals. The A4 Study is funded by a public-private-philanthropic partnership, including funding from the National Institutes of Health-National Institute on Aging, Eli Lilly and Company, Alzheimer’s Association, Accelerating Medicines Partnership, GHR Foundation, an anonymous foundation and additional private donors, with in-kind support from Avid and Cogstate. The companion observational Longitudinal Evaluation of Amyloid Risk and Neurodegeneration (LEARN) Study is funded by the Alzheimer’s Association and GHR Foundation. The A4 and LEARN Studies are led by Dr. Reisa Sperling at Brigham and Women’s Hospital, Harvard Medical School and Dr. Paul Aisen at the Alzheimer’s Therapeutic Research Institute (ATRI), University of Southern California. The A4 and LEARN Studies are coordinated by ATRI at the University of Southern California, and the data are made available through the Laboratory for Neuro Imaging at the University of Southern California. The participants screening for the A4 Study provided permission to share their de-identified data in order to advance the quest to find a successful treatment for Alzheimer’s disease. We would like to acknowledge the dedication of all the participants, the site personnel, and all of the partnership team members who continue to make the A4 and LEARN Studies possible. The complete A4 Study Team list is available on: a4study.org/a4-study-team.; the Adult Changes in Thought study (ACT), U01 AG006781, U19 AG066567; Alzheimer’s Disease Neuroimaging Initiative (ADNI): Data collection and sharing for this project was funded by the Alzheimer’s Disease Neuroimaging Initiative (ADNI) (National Institutes of Health Grant U01 AG024904) and DOD ADNI (Department of Defense award number W81XWH-12-2-0012). ADNI is funded by the National Institute on Aging, the National Institute of Biomedical Imaging and Bioengineering, and through generous contributions from the following: AbbVie, Alzheimer’s Association; Alzheimer’s Drug Discovery Foundation; Araclon Biotech; BioClinica, Inc.; Biogen; Bristol-Myers Squibb Company; CereSpir, Inc.; Cogstate; Eisai Inc.; Elan Pharmaceuticals, Inc.; Eli Lilly and Company; EuroImmun; F. Hoffmann-La Roche Ltd and its affiliated company Genentech, Inc.; Fujirebio; GE Healthcare; IXICO Ltd.;Janssen Alzheimer Immunotherapy Research & Development, LLC.; Johnson & Johnson Pharmaceutical Research & Development LLC.; Lumosity; Lundbeck; Merck & Co., Inc.;Meso Scale Diagnostics, LLC.; NeuroRx Research; Neurotrack Technologies; Novartis Pharmaceuticals Corporation; Pfizer Inc.; Piramal Imaging; Servier; Takeda Pharmaceutical Company; and Transition Therapeutics. The Canadian Institutes of Health Research is providing funds to support ADNI clinical sites in Canada. Private sector contributions are facilitated by the Foundation for the National Institutes of Health (www.fnih.org). The grantee organization is the Northern California Institute for Research and Education, and the study is coordinated by the Alzheimer’s Therapeutic Research Institute at the University of Southern California. ADNI data are disseminated by the Laboratory for Neuro Imaging at the University of Southern California; Estudio Familiar de Influencia Genetica en Alzheimer (EFIGA): 5R37AG015473, RF1AG015473, R56AG051876; Memory & Aging Project at Knight Alzheimer’s Disease Research Center (MAP at Knight ADRC): The Memory and Aging Project at the Knight-ADRC (Knight-ADRC). This work was supported by the National Institutes of Health (NIH) grants R01AG064614, R01AG044546, RF1AG053303, RF1AG058501, U01AG058922 and R01AG064877 to Carlos Cruchaga. The recruitment and clinical characterization of research participants at Washington University was supported by NIH grants P30AG066444, P01AG03991, and P01AG026276. Data collection and sharing for this project was supported by NIH grants RF1AG054080, P30AG066462, R01AG064614 and U01AG052410. We thank the contributors who collected samples used in this study, as well as patients and their families, whose help and participation made this work possible. This work was supported by access to equipment made possible by the Hope Center for Neurological Disorders, the Neurogenomics and Informatics Center (NGI: https://neurogenomics.wustl.edu/) and the Departments of Neurology and Psychiatry at Washington University School of Medicine; National Alzheimer’s Coordinating Center (NACC): The NACC database is funded by NIA/NIH Grant U24 AG072122. NACC data are contributed by the NIA-funded ADRCs: P30 AG062429 (PI James Brewer, MD, PhD), P30 AG066468 (PI Oscar Lopez, MD), P30 AG062421 (PI Bradley Hyman, MD, PhD), P30 AG066509 (PI Thomas Grabowski, MD), P30 AG066514 (PI Mary Sano, PhD), P30 AG066530 (PI Helena Chui, MD), P30 AG066507 (PI Marilyn Albert, PhD), P30 AG066444 (PI John Morris, MD), P30 AG066518 (PI Jeffrey Kaye, MD), P30 AG066512 (PI Thomas Wisniewski, MD), P30 AG066462 (PI Scott Small, MD), P30 AG072979 (PI David Wolk, MD), P30 AG072972 (PI Charles DeCarli, MD), P30 AG072976 (PI Andrew Saykin, PsyD), P30 AG072975 (PI David Bennett, MD), P30 AG072978 (PI Neil Kowall, MD), P30 AG072977 (PI Robert Vassar, PhD), P30 AG066519 (PI Frank LaFerla, PhD), P30 AG062677 (PI Ronald Petersen, MD, PhD), P30 AG079280 (PI Eric Reiman, MD), P30 AG062422 (PI Gil Rabinovici, MD), P30 AG066511 (PI Allan Levey, MD, PhD), P30 AG072946 (PI Linda Van Eldik, PhD), P30 AG062715 (PI Sanjay Asthana, MD, FRCP), P30 AG072973 (PI Russell Swerdlow, MD), P30 AG066506 (PI Todd Golde, MD, PhD), P30 AG066508 (PI Stephen Strittmatter, MD, PhD), P30 AG066515 (PI Victor Henderson, MD, MS), P30 AG072947 (PI Suzanne Craft, PhD), P30 AG072931 (PI Henry Paulson, MD, PhD), P30 AG066546 (PI Sudha Seshadri, MD), P20 AG068024 (PI Erik Roberson, MD, PhD), P20 AG068053 (PI Justin Miller, PhD), P20 AG068077 (PI Gary Rosenberg, MD), P20 AG068082 (PI Angela Jefferson, PhD), P30 AG072958 (PI Heather Whitson, MD), P30 AG072959 (PI James Leverenz, MD); National Institute on Aging Alzheimer’s Disease Family Based Study (NIA-AD FBS): U24 AG056270; Religious Orders Study (ROS): P30AG10161,R01AG15819, R01AG42210; Memory and Aging Project (MAP -Rush): R01AG017917, R01AG42210; Minority Aging Research Study (MARS): R01AG22018, R01AG42210; Washington Heights/Inwood Columbia Aging Project (WHICAP): RF1 AG054023;and Wisconsin Registry for Alzheimer’s Prevention (WRAP): R01AG027161 and R01AG054047. Additional acknowledgments include the National Institute on Aging Genetics of Alzheimer’s Disease Data Storage Site (NIAGADS, U24AG041689) at the University of Pennsylvania, funded by NIA.

## Consent Statement

Consent from human subjects was not necessary for this study.

